# The association of C-Reactive Protein with Delirium Severity: An analysis of a prospective cohort study

**DOI:** 10.64898/2026.10.22.26366074

**Authors:** Fiona Schnitzler, Kaitlin Kramer, Samantha Curtis, Belinda Anderson, Kyriakos Matsis, Amy Lawrence, Gerald Wong, Benjamin Moran, Neil Pillinger, Robert D. Sanders

## Abstract

**Background:** Postoperative delirium (POD) is common in elderly patients and is linked to poor recovery and greater mortality. Inflammation has been implicated in its pathophysiology and recent meta-analysis suggests steroids may have utility in preventing POD. This study aimed to evaluate whether C-reactive protein (CRP) predicts severity (or incidence) of POD. Our secondary outcome analysed whether anti-emetic doses of dexamethasone meaningfully altered inflammatory responses in this age group.

**Methods:** Patients (n=122) ≥60 years of age undergoing major surgery were recruited to a prospective cohort study in which CRP was measured preoperatively and on postoperative day 1. Delirium was assessed using the 3-minute diagnostic Confusion Assessment Method (3D-CAM) twice daily on days 1–3. CRP was analysed against the outcomes of incidence (positive overall 3D-CAM) and peak severity (maximum 3D-CAM score) of POD.

**Results:** POD occurred in 20/122 (16.4%); mean peak severity 2.1 (SD 2.1). Postoperative CRP was associated with the primary outcome, POD severity (predicted probability 94.7%) as well as incidence (predicted probability 99.9%). Dexamethasone reduced CRP by 41% (95% CrI 8-62%, predicted probability 99.1%).

**Conclusion:** Routine postoperative CRP was independently associated with POD incidence and severity. Dexamethasone, at anti-emetic doses, was associated with a substantial reduction in CRP levels, supporting trials focussed on modulating inflammation to prevent delirium.

---

Delirium is an acute neurocognitive disorder, predominantly affecting older people, characterised by an acute, fluctuating decline in attention, cognition, and arousal. Delirium occurs frequently in the postoperative period (11-51%), often referred to as the most common surgical complication.^1^ While delirium itself is transient, the consequences are considerable and persisting. Delirium has been associated with longer hospital stay, 4-fold 30-day readmission risk,^2, 3^ and a 3-7-fold greater risk of mortality.^3-5^ The risks associated with delirium are not always immediately manifest however, and there is a growing body of research reflecting an insidious course of functional and cognitive decline following delirium.^5-8^ Annual healthcare costs due to delirium have been estimated at $152 billion in the USA alone, with daily costs being nearly 3-fold greater for those with vs without POD.^9^

It is widely recognised that inflammation is a core feature of the pathophysiology of POD, wherein it may act to increase permeability of the blood brain barrier, perturb cellular energetics and synaptic activity, and disturb information transfer in the brain.^10-13^ While various cytokines have been linked to POD, it is unclear how useful routinely collected inflammatory markers such as C-Reactive Protein (CRP) or white cell counts may be.^14^ Establishing this link would be useful for multicenter trials of anti-inflammatory therapies, where routine markers may help gauge the pathophysiological impact of the therapy. This would be particularly important in establishing a causal link between reducing inflammation and consequent effects on delirium.

The current utility of preoperative CRP is unclear, with some studies demonstrating a link to the incidence,^15-18^ severity and duration^15^ of POD, while another showed no association.^19^ High postoperative CRP has been well linked to incidences of POD^15, 20-24^ but few studies have examined duration and severity.^15^ Interpretation is further limited as CRP rises non-specifically after surgery, so it remains unclear how closely it relates to delirium once clinical factors such as duration of surgery are considered. Validating a dose-dependent relationship between CRP and delirium severity would enhance biological plausibility that modulating CRP may meaningfully alter delirium.

A recent meta-analysis suggested potential utility of perioperative corticosteroids to reduce POD but concerns over risk of bias and generalisabilty meant that recommendations for clinical practice were not issued.^25^ A concern from the literature is in determining the most effective dose of corticosteroid for reducing inflammation – with doses from 0.6mg/kg to 1mg/kg dexamethasone reducing inflammation by ∼40%.^26, 27^ This implies they may have already reached a ceiling in the anti-inflammatory effects of dexamethasone. Whether anti-emetic dosing may also reduce inflammatory markers to a similar extent is therefore unknown.

Given the limited evidence for a dose-dependent effect of CRP on delirium severity, our primary objective was to investigate the association between postoperative CRP and peak delirium severity. Establishing dose dependence is a key factor in determining the rationale for anti-inflammatory drugs in the prevention and treatment of delirium. Secondary objectives were to investigate the association between CRP and the incidence of delirium, including after adjustment for operative factors such as length of surgery, and to examine demographic and operative risk factors for delirium. An exploratory analysis of intraoperative dexamethasone administration sought to identify associations with postoperative CRP. We hypothesised that postoperative CRP would be positively correlated with peak delirium severity and incidence of delirium, and that dexamethasone would reduce postoperative CRP, providing rationale for a trial of dexamethasone to prevent delirium.

## Methods

Improving Perioperative Outcomes after Surgical Events (IMPOSE) was a prospective longitudinal cohort study with 122 participants. Participants were recruited if they met the inclusion criteria of undergoing a major surgery (requiring at least a 2-day hospital stay) at Royal Prince Alfred Hospital and were ≥60 years old at the time of surgery. This study received ethics approval in June 2023 (Protocol No. X23-0165 & 2023/ETH00923).

Demographic data were collected preoperatively via patient account and the electronic Medical Record (eMR). Delirium assessments were performed using the 3-minute diagnostic Confusion Assessment Method (3D-CAM)^9^ or CAM-ICU in ventilated patients^29^ once during the preoperative visit, and in the morning and afternoon on postoperative days 1-3, depending on length of stay. Data were deidentified and securely stored in REDCap. Baseline cognitive function was assessed using the Controlled Oral Word Association Test (COWAT).^28^ The American College of Surgeons National Surgical Quality Improvement Program (NSQIP) Surgical Risk Calculator^30^ was used to predict risk of 30-day mortality for each patient with information gathered from the eMR. CRP was measured using the Nucleic acid Linked Immuno-Sandwich Assay (NULISA) by Geneworks (Melbourne, Australia).^31^ Blood sample data were collected preoperatively and on postoperative day 1. Hospital CRP levels were collected from the eMR and were used to compare with NULISA assay levels. Intraoperative steroid exposure with anti-emetic doses of dexamethasone was collected from the eMR.

### Statistical Analysis

Analyses were conducted in R 3.19.0^32^ using brms 2.23.0^33^ as an interface to CmdStan 2.39.0. Models were run on a 64-bit Linux system (Ubuntu 22.04.5 LTS). All models used brms default priors: improper flat priors on regression coefficients, weakly informative Student-t priors on the intercept and, for linear models, the residual standard deviation, and an LKJ(1) prior on correlations. Each model was run with 4 chains of 4,000 iterations (1,000 warm-up), and convergence was confirmed by 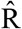< 1.01 for all parameters. Analyses used complete cases. Estimates are reported with 95% credible intervals (CrI), alongside the posterior probability that the effect lies in the stated direction (e.g. P(OR > 1), the probability that the odds of delirium are increased).

Peak delirium severity (0-21) was calculated by the maximum sum of positive items on the 3D-CAM at a single postoperative time point for each patient. Baseline cognitive function was calculated by the sum of correct words recalled in two COWATs. Continuous covariates were rescaled so that effects are reported per 10 years of age, per standard deviation (SD) of NSQIP risk and baseline cognition, and per hour of surgery.

CRP was measured using the NULISA assay and reported in NULISA Protein Quantification (NPQ) units (log2 scale).^31^ The correlation between hospital laboratory CRP and NULISA CRP was estimated separately for preoperative and postoperative samples. Hospital CRP was log2-transformed to reduce skewness; NULISA CRP values (NPQ) are on a log2 scale. Both measures were standardised, and a Bayesian bivariate normal model was fitted using patients with both measurements available (preoperative n=106; postoperative n=88). The correlation coefficient (r) is reported as the posterior median with 95% credible interval. Similarly, NULISA CRP was correlated with length of surgery, given the assumption that surgical severity is the primary driver of the postoperative inflammatory response.

Associations of pre- and postoperative NULISA CRP with delirium incidence and severity were estimated using Bayesian logistic and linear regression, respectively, with both time points entered in the same model so that each estimate is adjusted for the other. Three models were fitted: (1) unadjusted; (2) primary model: adjusted for age, sex, cardiac surgery, baseline cognitive function and NSQIP 30- day mortality risk; and (3) additionally adjusted for length of surgery.

In a secondary analysis restricted to non-cardiac surgery patients with known steroid status, the association of intraoperative dexamethasone with postoperative CRP was estimated using Bayesian linear regression, and its association with delirium incidence using Bayesian logistic regression. For CRP, dexamethasone (any dose vs none) was examined in models fitted unadjusted, adjusted for preoperative CRP, and additionally adjusted for age, sex, NSQIP 30-day mortality risk and length of surgery. Dose was examined in the fully adjusted model by categorising dexamethasone as none, 4 mg or 8 mg; one patient who received 12 mg was excluded from this analysis. Because NULISA CRP is on a log2 scale, effects on CRP were expressed as percentage change, calculated as (2^β − 1) × 100. Delirium models (any dose vs none) were fitted unadjusted, adjusted for age, sex, NSQIP 30-day mortality risk and baseline cognitive function, and additionally adjusted for length of surgery.

## Results

Cohort characteristics are presented in Table 1. Delirium occurred in 20 of 122 patients (16.4%), and 93 (76.9%) scored positively on at least 1 item of the 3D-CAM. Two of these patients were identified as delirious using the CAM-ICU. Median peak delirium severity was 2 (IQR 1-3) overall and 5 (IQR 3-7) in patients with delirium (Supplementary Figure 1). Patients who developed delirium (n=20) had a higher NSQIP 30-day mortality risk, longer operations, and were more likely to have undergone cardiac surgery or to have pre-existing atrial fibrillation (Table 1).

**Table 1.**
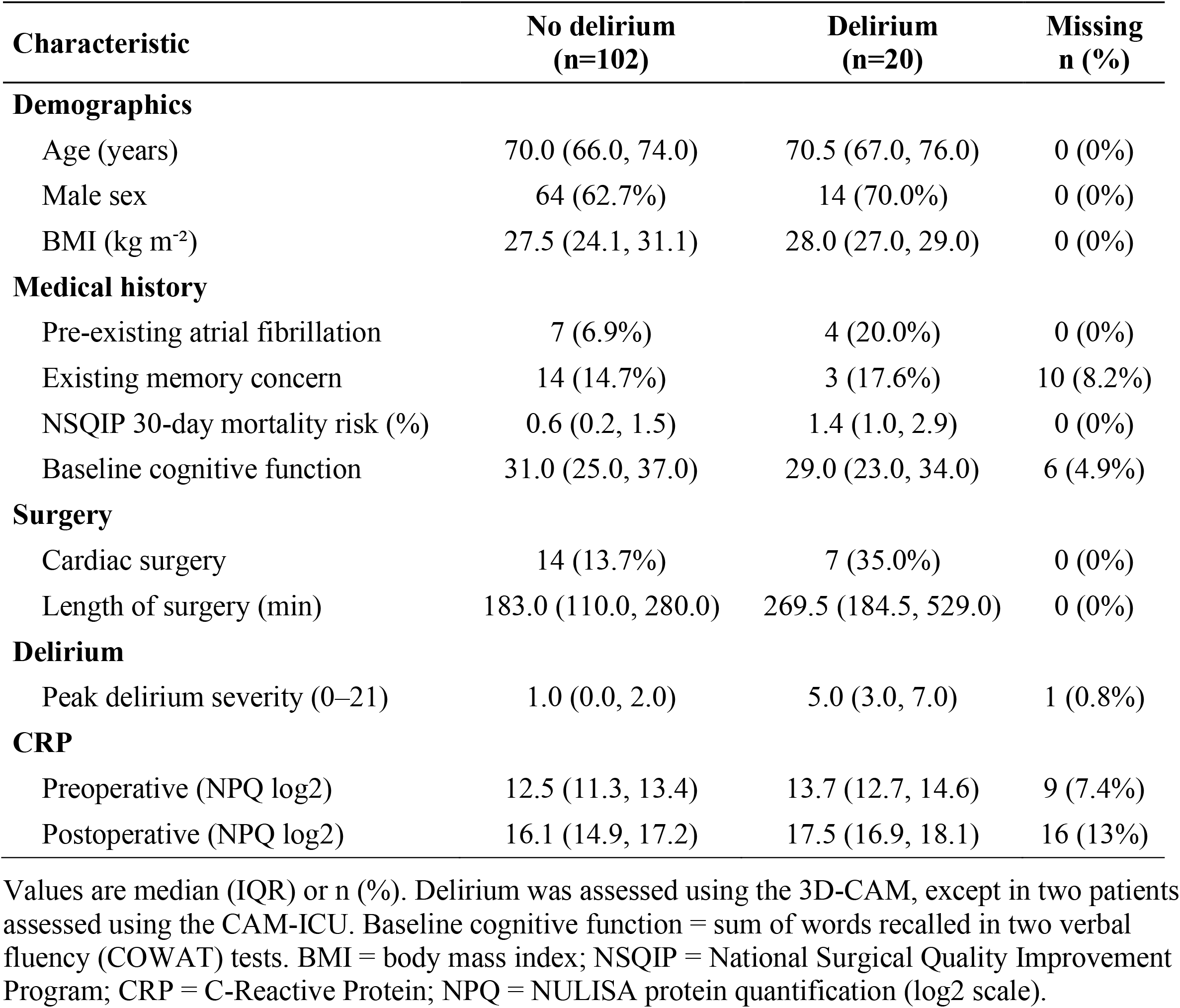
Characteristics of the cohort by postoperative delirium status.

NULISA CRP was strongly correlated with hospital CRP before surgery (r=0.93, 95% CrI [0.90, 0.95]) and postoperatively (r=0.93, [0.89, 0.95], supplementary Figure 2). Median NULISA CRP was numerically higher in patients who developed delirium than in those who did not, both preoperatively (13.7 vs 12.5 NPQ) and postoperatively (17.5 vs 16.1 NPQ), corresponding to approximately 2.3-fold and 2.6-fold higher CRP, respectively (Table 1 and supplementary Figure 3).

### Peak delirium severity

For the primary outcome, postoperative CRP was positively associated with delirium severity in the unadjusted model (0.26 severity points per doubling of CRP, 95% CrI [0.03, 0.50]; P(>0) = 0.98; n = 100) and, with less certainty, after adjustment for age, sex, cardiac surgery, baseline cognitive function and NSQIP 30-day mortality risk (0.23, [−0.05, 0.51]; P(>0) = 0.95; n = 94) (Table 2 and Figure 2). This association was markedly attenuated after additional adjustment for length of surgery (0.07, [−0.23, 0.37]; P(>0) = 0.67), which was itself associated with higher severity (0.22 points per hour, [0.05, 0.38]; P(>0) = 0.99) (see Supplementary Figure 4 for correlations between CRP and length of surgery).

**Table 2.**
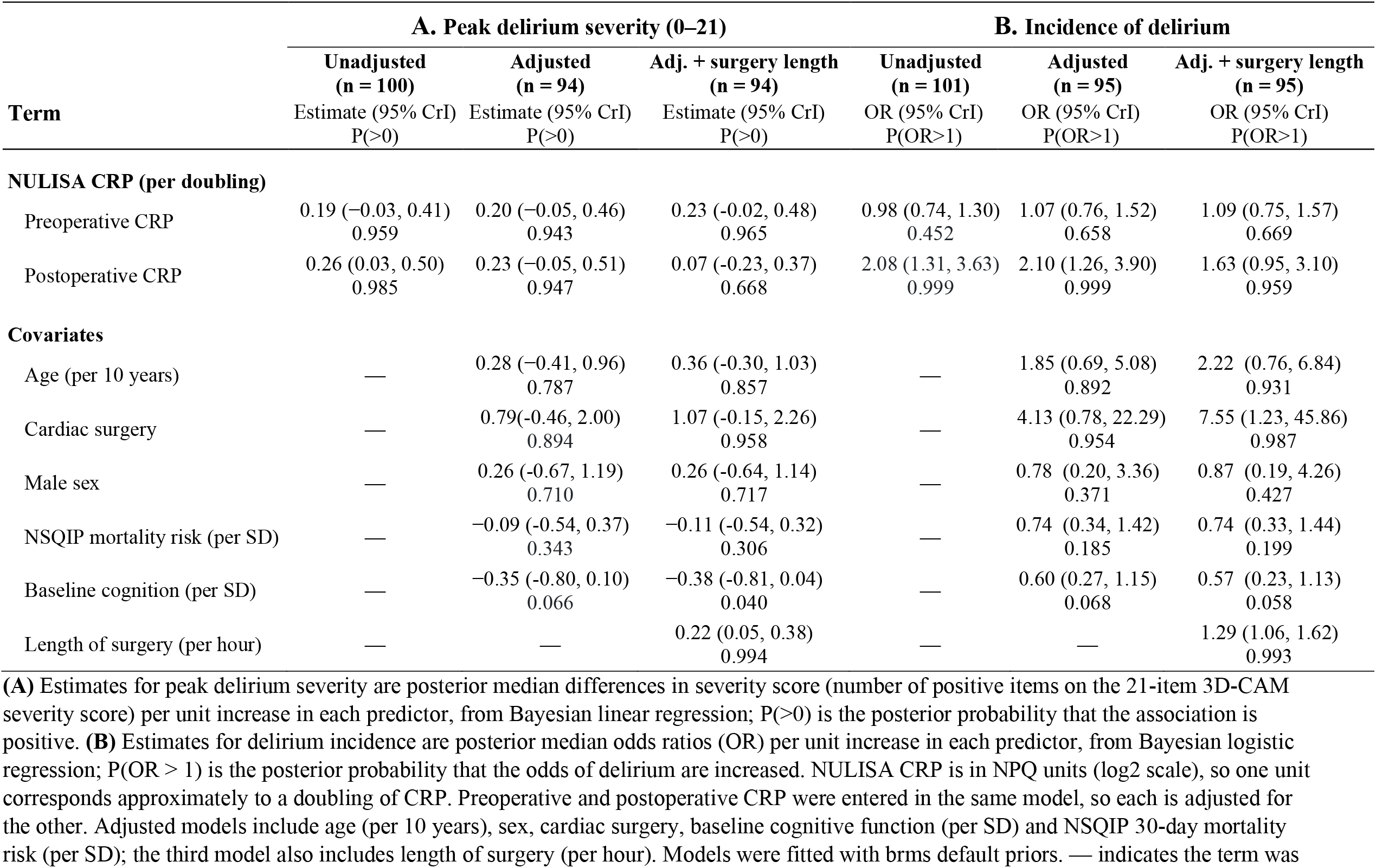
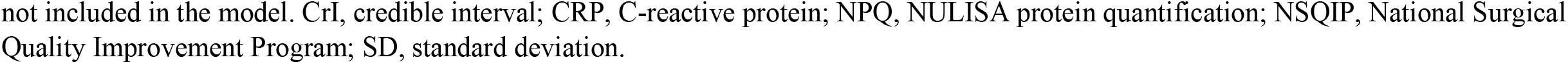
Association between NULISA CRP and **(A)** peak delirium severity and **(B)** incidence of delirium.

| Term | A. Peak delirium severity (0–21) |  |  | B. Incidence of delirium |  |  |
| --- | --- | --- | --- | --- | --- | --- |
|  | Unadjusted<br>(n = 100) | Adjusted<br>(n = 94) | Adj. + surgery length<br>(n = 94) | Unadjusted<br>(n = 101) | Adjusted<br>(n = 95) | Adj. + surgery length<br>(n = 95) |
|  | Estimate (95% CrI)<br>P(>0) | Estimate (95% CrI)<br>P(>0) | Estimate (95% CrI)<br>P(>0) | OR (95% CrI)<br>P(OR>1) | OR (95% CrI)<br>P(OR>1) | OR (95% CrI)<br>P(OR>1) |
| **NULISA CRP (per doubling)** |  |  |  |  |  |  |
| Preoperative CRP | 0.19 (−0.03, 0.41)<br>0.959 | 0.20 (−0.05, 0.46)<br>0.943 | 0.23 (−0.02, 0.48)<br>0.965 | 0.98 (0.74, 1.30)<br>0.452 | 1.07 (0.76, 1.52)<br>0.658 | 1.09 (0.75, 1.57)<br>0.669 |
| Postoperative CRP | 0.26 (0.03, 0.50)<br>0.985 | 0.23 (−0.05, 0.51)<br>0.947 | 0.07 (−0.23, 0.37)<br>0.668 | 2.08 (1.31, 3.63)<br>0.999 | 2.10 (1.26, 3.90)<br>0.999 | 1.63 (0.95, 3.10)<br>0.959 |
| **Covariates** |  |  |  |  |  |  |
| Age (per 10 years) | — | 0.28 (−0.41, 0.96)<br>0.787 | 0.36 (−0.30, 1.03)<br>0.857 | — | 1.85 (0.69, 5.08)<br>0.892 | 2.22 (0.76, 6.84)<br>0.931 |
| Cardiac surgery | — | 0.79(−0.46, 2.00)<br>0.894 | 1.07 (−0.15, 2.26)<br>0.958 | — | 4.13 (0.78, 22.29)<br>0.954 | 7.55 (1.23, 45.86)<br>0.987 |
| Male sex | — | 0.26 (−0.67, 1.19)<br>0.710 | 0.26 (−0.64, 1.14)<br>0.717 | — | 0.78 (0.20, 3.36)<br>0.371 | 0.87 (0.19, 4.26)<br>0.427 |
| NSQIP mortality risk (per SD) | — | −0.09 (−0.54, 0.37)<br>0.343 | −0.11 (−0.54, 0.32)<br>0.306 | — | 0.74 (0.34, 1.42)<br>0.185 | 0.74 (0.33, 1.44)<br>0.199 |
| Baseline cognition (per SD) | — | −0.35 (−0.80, 0.10)<br>0.066 | −0.38 (−0.81, 0.04)<br>0.040 | — | 0.60 (0.27, 1.15)<br>0.068 | 0.57 (0.23, 1.13)<br>0.058 |
| Length of surgery (per hour) | — | — | 0.22 (0.05, 0.38)<br>0.994 | — | — | 1.29 (1.06, 1.62)<br>0.993 |

(A) Estimates for peak delirium severity are posterior median differences in severity score (number of positive items on the 21-item 3D-CAM severity score) per unit increase in each predictor, from Bayesian linear regression; P(>0) is the posterior probability that the association is positive. (B) Estimates for delirium incidence are posterior median odds ratios (OR) per unit increase in each predictor, from Bayesian logistic regression; P(OR > 1) is the posterior probability that the odds of delirium are increased. NULISA CRP is in NPQ units (log2 scale), so one unit corresponds approximately to a doubling of CRP. Preoperative and postoperative CRP were entered in the same model, so each is adjusted for the other. Adjusted models include age (per 10 years), sex, cardiac surgery, baseline cognitive function (per SD) and NSQIP 30-day mortality risk (per SD); the third model also includes length of surgery (per hour). Models were fitted with brms default priors. — indicates the term was
not included in the model. CrI, credible interval; CRP, C-reactive protein; NPQ, NULISA protein quantification; NSQIP, National Surgical Quality Improvement Program; SD, standard deviation.

**Figure 1.**
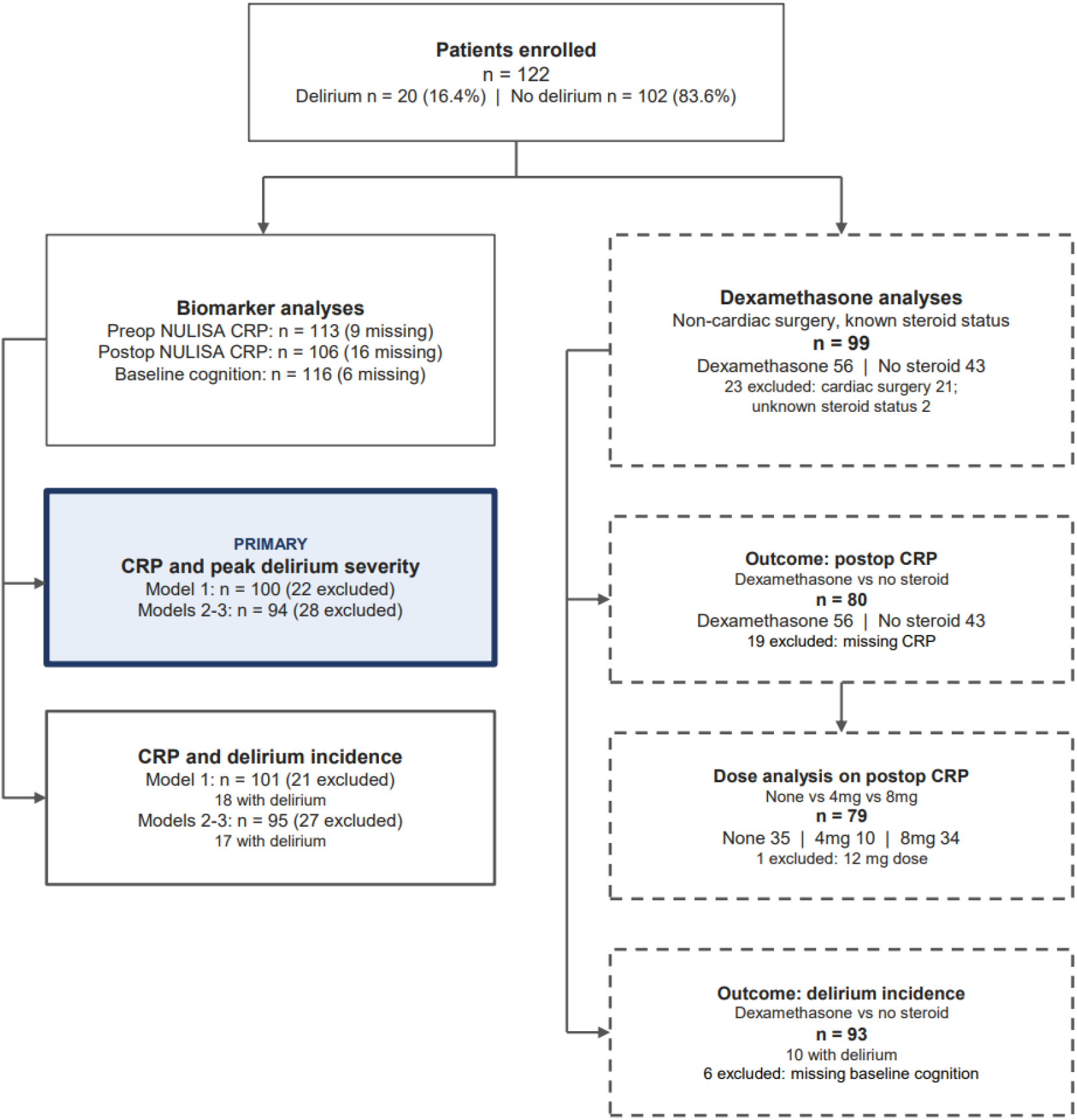
Participant flow diagram. Complete-case analysis. Excluded = missing data for any variable the model requires (pre- or postoperative CRP, severity score, and baseline cognition for adjusted models). Model 1 = unadjusted; Model 2 = adjusted for age, cardiac surgery, sex, NSQIP 30-day mortality risk and baseline cognition; Model 3 = additionally includes length of surgery. NULISA = Nucleic acid Linked Immuno-Sandwich Assay; CRP = C-Reactive Protein; NSQIP = National Surgical Quality Improvement Program.

**Figure 2.**
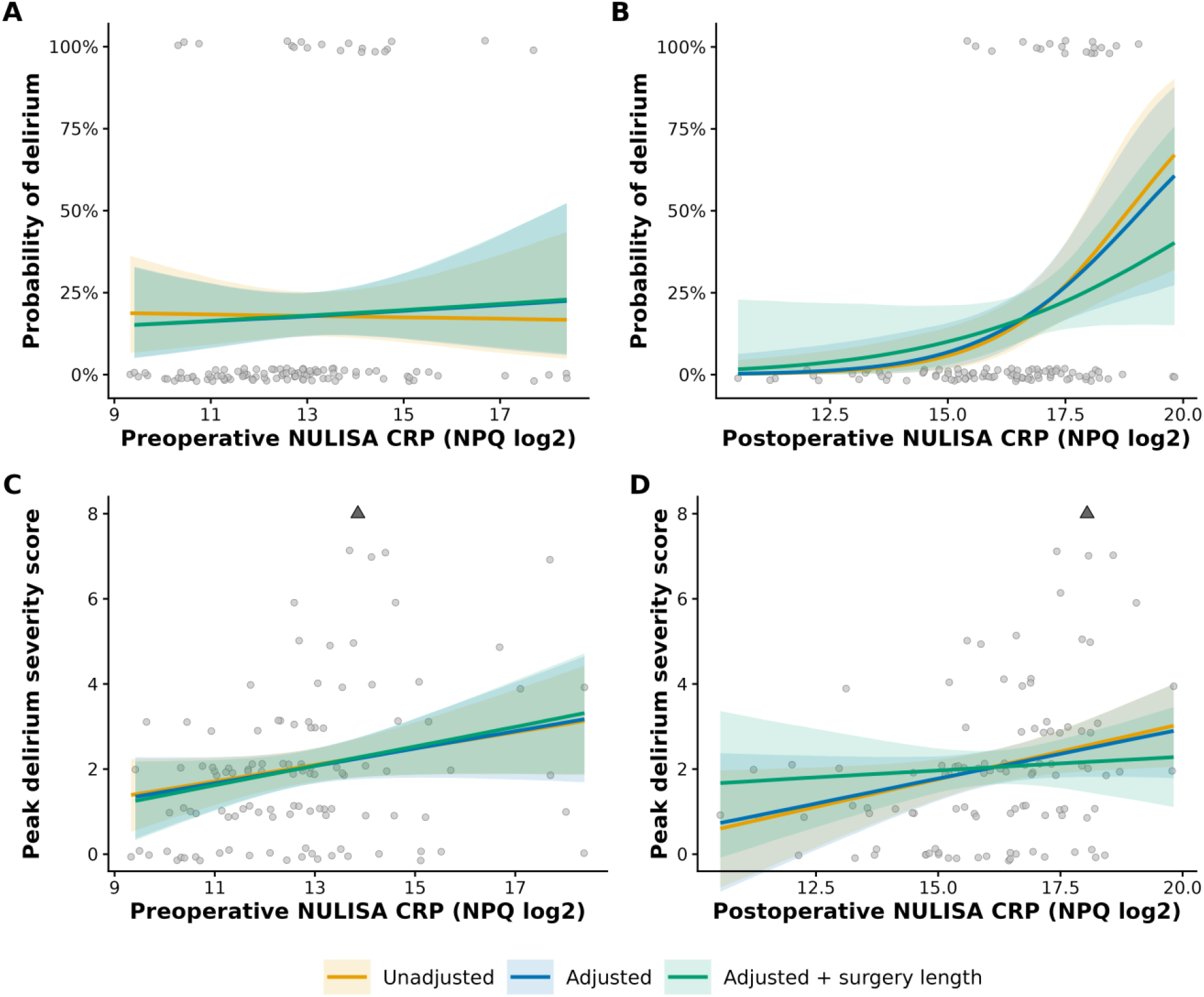
Predicted postoperative delirium and delirium severity by NULISA CRP. Population-averaged predicted probability of delirium (A and B; Bayesian logistic regression) and peak delirium severity score (C and D; Bayesian linear regression) across preoperative (A and C) and postoperative day 1 (B and D) NULISA CRP (NPQ log2 scale). Models were unadjusted; adjusted for age, sex, cardiac surgery, baseline cognitive function and NSQIP 30-day mortality risk; and additionally adjusted for length of surgery. Each model includes both preoperative and postoperative CRP. Lines show posterior medians and shaded bands 95% credible intervals. Grey points show observed patients (slight vertical jitter added); in A and B, points at 100% had delirium and points at 0% did not. One patient with a severity score of 14 lies above the plotted range in C and D and is shown as a triangle at the upper edge; this patient was included in all models. NULISA = Nucleic acid Linked Immuno-Sandwich Assay; CRP = C-reactive protein.

Preoperative CRP showed consistently positive estimates across all three models, although all 95% CrIs included zero (adjusted: 0.20, [−0.05, 0.46]; P(>0) = 0.94); unlike postoperative CRP, its estimate remained similar after adjustment for length of surgery (0.23, [−0.02, 0.48]; P(>0) = 0.97). Better baseline cognitive function was associated with lower severity (−0.35 points per SD in the adjusted model, [−0.80, 0.10]; P(<0) = 0.93).

### Delirium incidence

Each doubling of postoperative CRP was associated with higher odds of delirium in the unadjusted model (OR 2.08, 95% CrI [1.31, 3.63]; P(OR > 1) > 0.99; n = 101, 18 with delirium) and after adjustment for age, sex, cardiac surgery, baseline cognitive function and NSQIP 30-day mortality risk (OR 2.10, [1.26, 3.90]; P(OR > 1) > 0.99; n = 95, 17 with delirium) (Table 3 and Figure 2). This association was moderately attenuated after additional adjustment for length of surgery (OR 1.63, [0.95, 3.10]; P(OR > 1) = 0.96), which was itself associated with delirium (OR 1.29 per hour, [1.06, 1.62]; P(OR > 1) = 0.99). Preoperative CRP was not associated with delirium in any model (OR 0.98 to 1.09). Better baseline cognitive function was associated with lower odds of delirium in the adjusted model, although with some uncertainty (OR 0.60 per SD, [0.27, 1.15]; P(OR < 1) = 0.93).

**Table 3.** Effect of dexamethasone on postoperative NULISA CRP.

| Term | Unadjusted<br>(n = 80) |  | Adj. for preoperative CRP<br>(n = 80) |  | Adj. for preoperative CRP + covariates<br>(n = 80) |  |
| --- | --- | --- | --- | --- | --- | --- |
|  | Estimate (95% CrI)<br>P(>0) | % change<br>(95% CrI) | Estimate (95% CrI)<br>P(>0) | % change<br>(95% CrI) | Estimate (95% CrI)<br>P(>0) | % change<br>(95% CrI) |
| **Exposure** |  |  |  |  |  |  |
| Dexamethasone (vs none) | -0.84 (-1.68, -0.02)<br>0.024 | -44%<br>(-69%, -1%) | -0.54 (-1.22, 0.15)<br>0.066 | -31%<br>(-57%, 11%) | -0.76 (-1.40, -0.13)<br>0.009 | -41%<br>(-62%, -8%) |
| **Covariates** |  |  |  |  |  |  |
| Preoperative CRP (per doubling) | — | — | 0.50 (0.34, 0.67)<br>1.000 | 42%<br>(26%, 60%) | 0.42 (0.26, 0.58)<br>1.000 | 33%<br>(20%, 49%) |
| Age (per 10 years) | — | — | — | — | -0.40 (-0.89, 0.12)<br>0.064 | -24%<br>(-46%, 8%) |
| Male sex | — | — | — | — | 0.31 (-0.32, 0.94)<br>0.839 | 24%<br>(-20%, 91%) |
| NSQIP mortality risk (per SD) | — | — | — | — | 0.13 (-0.22, 0.49)<br>0.775 | 10%<br>(-14%, 40%) |
| Length of surgery (per hour) | — | — | — | — | 0.21 (0.12, 0.30)<br>1.000 | 15%<br>(8%, 23%) |

Estimates are posterior median differences in postoperative day 1 NULISA CRP per unit increase in each predictor, from Bayesian linear regression, shown on the NPQ (log2) scale and as percentage change in CRP, calculated as $(2^{\text{estimate}} - 1) \times 100$ ; P(>0) is the posterior probability of a positive association (higher CRP), so the probability that dexamethasone reduces CRP is $1 - P(>0)$ . The second model is adjusted for preoperative CRP (per NPQ unit, approximately a doubling of CRP); the third model additionally includes age (per 10 years), sex, NSQIP 30-day mortality risk (per SD) and length of surgery (per hour). All models included the same patients (n = 80; non-cardiac surgery patients with known steroid status and complete data). Models were fitted with brms default priors. — indicates the term was not included in the model. CrI, credible interval; CRP, C-reactive protein; NPQ, NULISA protein quantification; NSQIP, National Surgical Quality Improvement Program; SD, standard deviation.

### Impact of Dexamethasone

Of 99 non-cardiac surgery patients with known steroid status, 56 received dexamethasone (Supplementary Table 1); these patients had longer operations than those who did not (median 198 vs 170 min). Dexamethasone was associated with lower postoperative CRP in the fully adjusted model (−41%, 95% CrI [−62%, −8%]; P(reduction) = 0.99; n = 80) (Table 3). The association was weaker after adjustment for preoperative CRP alone (−31%, [−57%, 11%]; P(reduction) = 0.93) and strengthened with additional adjustment for length of surgery and other covariates, consistent with longer operations in patients receiving dexamethasone. Compared with no steroid, 8 mg was associated with 44% lower CRP ([−65%, −10%]; P(reduction) = 0.99), while the estimate for 4 mg was imprecise (−35%, [−67%, 30%]; P(reduction) = 0.89), and the two doses could not be distinguished (8 mg vs 4 mg: −14%, [−58%, 80%]; P(reduction) = 0.66; n = 79, one patient receiving 12 mg excluded) (Supplementary Table 2). Delirium occurred in 3 of 52 dexamethasone patients (5.8%) and 7 of 41 patients without steroid (17.1%) with complete data. After adjustment for age, sex, NSQIP 30-day mortality risk and baseline cognitive function, dexamethasone was associated with lower odds of delirium, although with considerable uncertainty given the small number of events (OR 0.29, [0.06, 1.20]; P(OR<1) = 0.96, n = 93, 10 with delirium) (Supplementary Table 3).

## Discussion

In this cohort of 122 adults undergoing major surgery, postoperative CRP was associated with a modest increase in peak delirium severity. Each doubling of CRP was associated with approximately two-fold odds of POD. Both associations weakened after accounting for length of surgery, which was itself associated with both outcomes, consistent with length of surgery being a driver of inflammation and delirium (with some information not captured by CRP, conveyed by length of surgery). In an exploratory analysis of non-cardiac patients, intraoperative dexamethasone was associated with lower postoperative CRP.

Consistent with prior knowledge, patients who experienced delirium were more likely to have lower baseline cognitive function and undergo longer surgery or cardiac surgery. The data suggest a possible correlation with older age, although this finding was imprecise. This may be due to the age distribution of this cohort, with 75% of patients being 75 or younger, and the relatively low incidence of POD. Of the operative risk factors, length of surgery demonstrated the highest likelihood of association with POD. Previous research has demonstrated a 6% increase in risk of delirium per 30 minutes of surgery time when correcting for demographic risk factors.^34^ This association may be due to the increasing degree of surgical insult, and the consequent magnitude of inflammation.^35, 36^ Cardiac surgery was also a more common factor in those with POD than without, although this estimate was imprecise, likely due to small sample size.

The primary expected outcome of this study was the association between CRP and peak delirium severity. Peak delirium severity was positively associated with higher postoperative CRP. This was less certain after adjustment for covariates, and the estimate was attenuated after correcting for length of surgery. This is consistent with the premise that the extent of surgical trauma is proportional to the resultant degree of inflammation.^35, 36^ Preoperative CRP was also associated with a slightly higher delirium severity score, although to a lesser degree than postoperative. The extent to which delirium severity was associated with CRP, while positive, was modest. This may be due to the narrow, right-skewed distribution of severity scores in the cohort, with only one patient scoring above 7 severity points on any assessment.

The utility of preoperative CRP may be limited, as this study found only a slight increase in delirium severity with increasing CRP, and no association with incidence of delirium. While one study failed to demonstrate an association between preoperative CRP and POD,^19^ others have.^15-18^ Overall, our data are consistent with much of the literature, though we think it unlikely that preoperative CRP will have much clinical utility in predicting POD.

The secondary outcome of the relationship between postoperative CRP and the incidence of delirium yielded clear results, wherein odds of delirium approximately doubled with each doubling of postoperative CRP. This supports previously established associations between postoperative CRP and delirium.^15, 16, 20-24^ While postoperative CRP increased odds of delirium in all models, adjustment for length of surgery diminished this finding – likely due to the intrinsic link between duration of surgery and degree of postoperative inflammation.^35, 36^ Longer procedures conferred greater risk of delirium, independent of CRP.

In non-cardiac surgical patients, intraoperative dexamethasone administration was associated with a lower postoperative CRP relative to patients who did not receive steroid. This effect was seen in both the adjusted and unadjusted models; but certainty was reduced when correcting for preoperative CRP. This may reflect the lower mean preoperative CRP in the dexamethasone compared to the no steroid group. 8mg dosing was credibly associated with lower postoperative CRP. The estimated reduction in CRP following 4mg dosing was less precise, as was the estimated superiority of 8mg over 4mg. This was likely a result of only 10 patients receiving 4mg. While we recommend caution in interpreting these results, dexamethasone administration was associated with a reduced incidence of delirium. Patients who received any dose of steroid had 72% lower odds of POD in both the adjusted and unadjusted models compared to patients who did not receive a dose. Estimated reduction in odds was greater following correction for length of surgery, however small sample sizes limit interpretation. Despite potentially inflated estimates, these data offer the hypothesis that dexamethasone may reduce incidence of POD, particularly in offsetting the inflammatory insult of longer surgery. Previous studies have demonstrated that pre- or intraoperative dosing with corticosteroids may reduce both incidence^37, 38^ and severity^37, 39^ of delirium. One systematic review found that severity of delirium was reduced and odds of delirium were approximately halved following administration of corticosteroids.^25^ In a non-cardiac cohort, a retrospective study of 92,832 patients found that rates of POD were lower in patients who received intraoperative dexamethasone relative to those who did not.^40^ However, it is worth emphasizing that the authors suggested a high quality randomized controlled trial was still required to establish an evidence base to support implementation of corticosteroids to prevent delirium.

Validation of the NULISA assay against hospital CRP also facilitates retrospective analysis of banked blood biomarkers to understand mechanisms of action of therapies that are nested within clinical trials.

Thus, clinical trials could attempt to either use hospital assays of CRP or store blood and analyse those samples post-hoc by NULISA. This is important, as if steroids modulate inflammation but not delirium, this would lead to a major re-appraisal of inflammatory models of delirium.

The main limitations of this study were the size of the cohort, and the incidence and distribution of delirium and operative factors. Analyses were performed using default priors on complete cases, restricting the cohort to 93-100 patients per model. The relatively low rate of delirium observed in this study reduced the precision of certain estimates, particularly in adjusted models and for dexamethasone analyses. Imprecision in associations between CRP and peak delirium severity is particularly plausible given the restricted range and right-skew of severity scores. Results of dexamethasone analyses were especially weak given small sample size, retrospective inclusion and unknown indications. Additionally, small cohort size precluded analyses of dexamethasone administration in cardiac surgical patients. We note the effect size for the dexamethasone effect is large and we consider this as the conflation of the biological effects of the drug and potentially selection bias. Regarding the latter point, it is interesting that surgeries appeared longer in the dexamethasone group.

Future research involving a larger cohort with a higher incidence of delirium, and well distributed peak delirium severity scores and demographic factors, may clarify the association between surgical inflammation and POD, especially relating to severity and duration. This may guide perioperative planning, including preoperative optimisation and intraoperative factors. A trial investigating the utility of intraoperative corticosteroids in mitigating postoperative inflammation may support these preventative measures.

## Conclusions

Postoperative CRP was associated with higher odds of delirium, and to a lesser extent, increased severity of delirium. These estimates were reduced when considering length of surgery, which was itself independently associated with both incidence and severity of POD. Intraoperative dexamethasone was associated with lower postoperative CRP in this exploratory analysis, which may suggest a potential to mitigate operative inflammation and potentially the burden of POD. These data are currently hypothesis-generating and warrant further evaluation in a larger prospective trial.

## Data Availability

Data may be available on request if consistent with ethics guidance.

