## Supplemental Figures and Tables for "The association of C-Reactive Protein with Delirium Severity: An analysis of a prospective cohort study"

### Supplementary Figures and Tables

**Supplementary Figure 1.** Distribution of peak delirium severity scores across cohort.

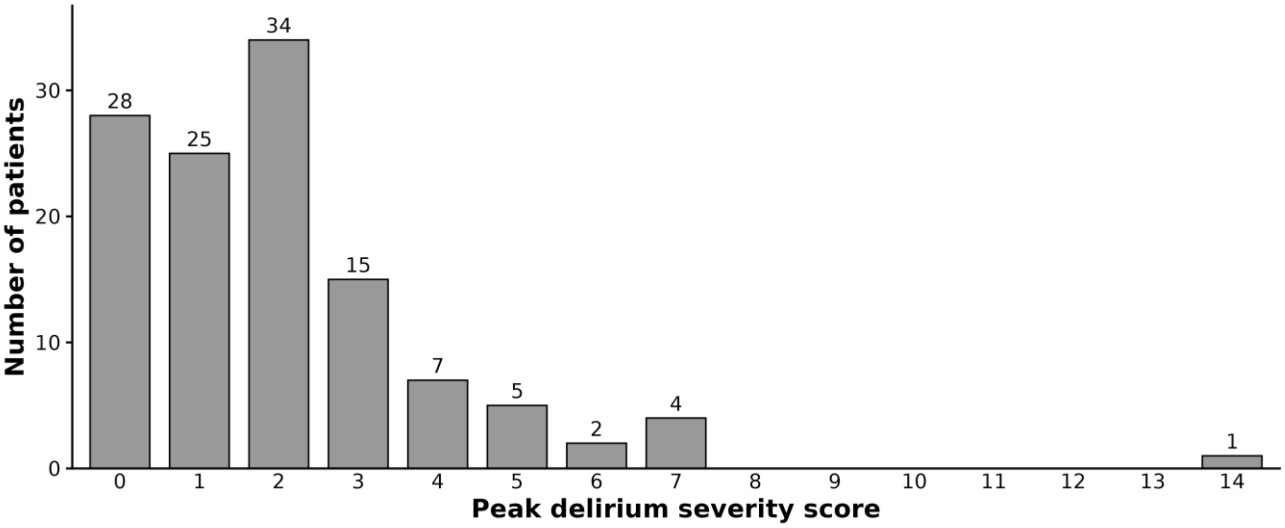


Peak delirium severity was calculated per patient as the highest number of positive items on the 3D-CAM on any one assessment, scored out of 21; n=121.

**Supplementary Figure 2.** Correlation between hospital and NULISA CRP.

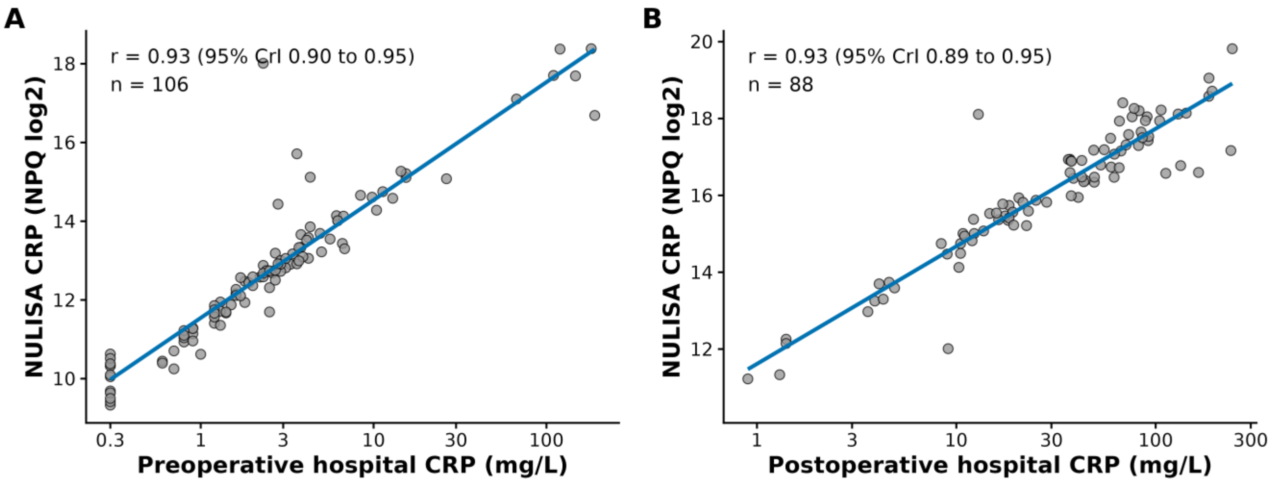


**(A)** Preoperative and **(B)** postoperative day 1 samples. Hospital CRP (mg/L) is shown on a logarithmic axis; NULISA CRP is shown in NPQ units, which are on a log2 scale. Each point represents one patient; the line shows the linear fit. r is the posterior median correlation between log2-transformed hospital CRP and NULISA CRP, estimated with a Bayesian bivariate normal model, with 95% credible interval (CrI). Hospital CRP values below the assay detection limit were recorded at the limit (0.3 mg/L). CRP = C-reactive protein; CrI = credible interval; NULISA = Nucleic acid Linked Immuno-Sandwich Assay; NPQ = NULISA protein quantification (log2 scale).

**Supplementary Figure 3.** Distributions of CRP.

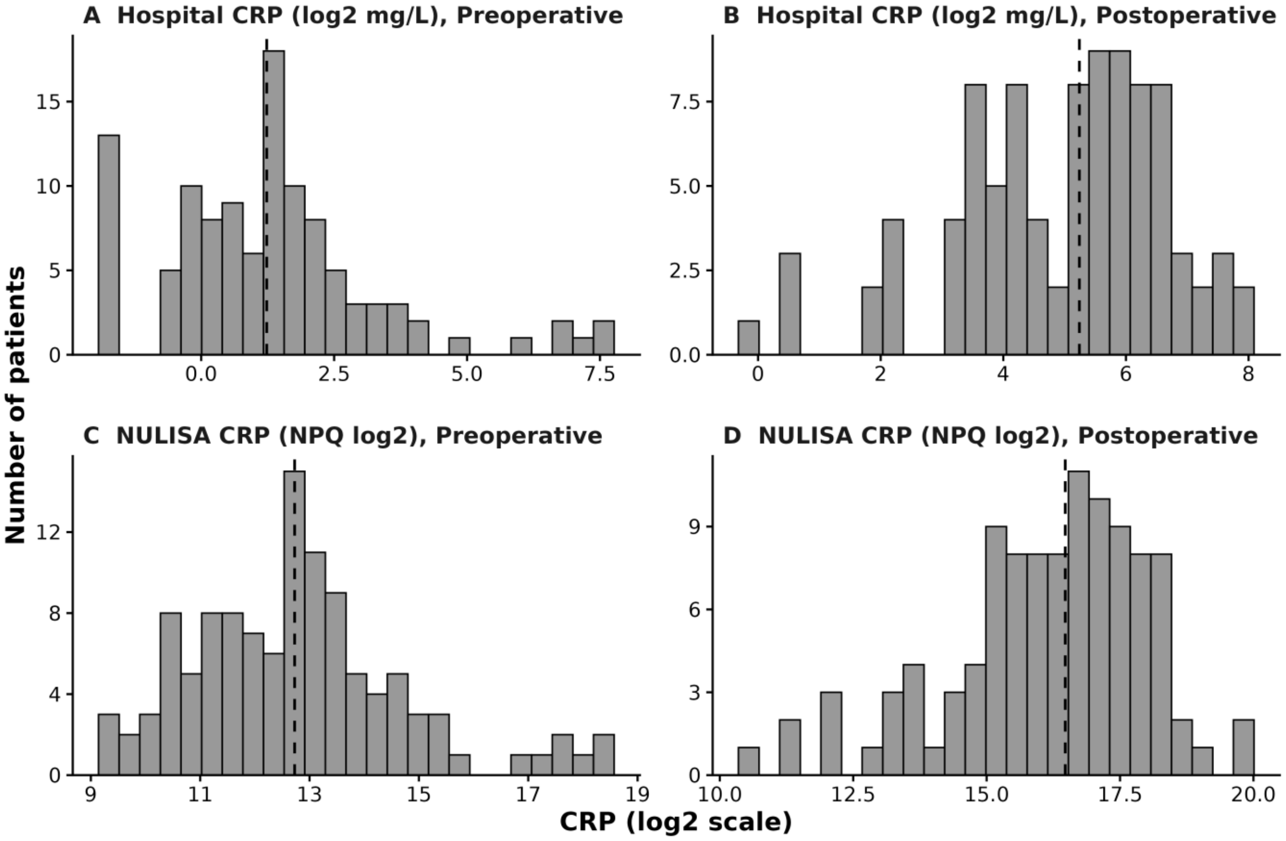


Hospital laboratory CRP (log2-transformed, mg/L) measured **(A)** preoperatively and **(B)** postoperative day 1, and NULISA CRP (NQP, log2 scale) measured **(C)** preoperatively and **(D)** on postoperative day 1. Dashed lines indicate medians. Each panel has its own axis scale.

**Supplementary Figure 4.** Correlation between NULISA CRP and length of surgery.

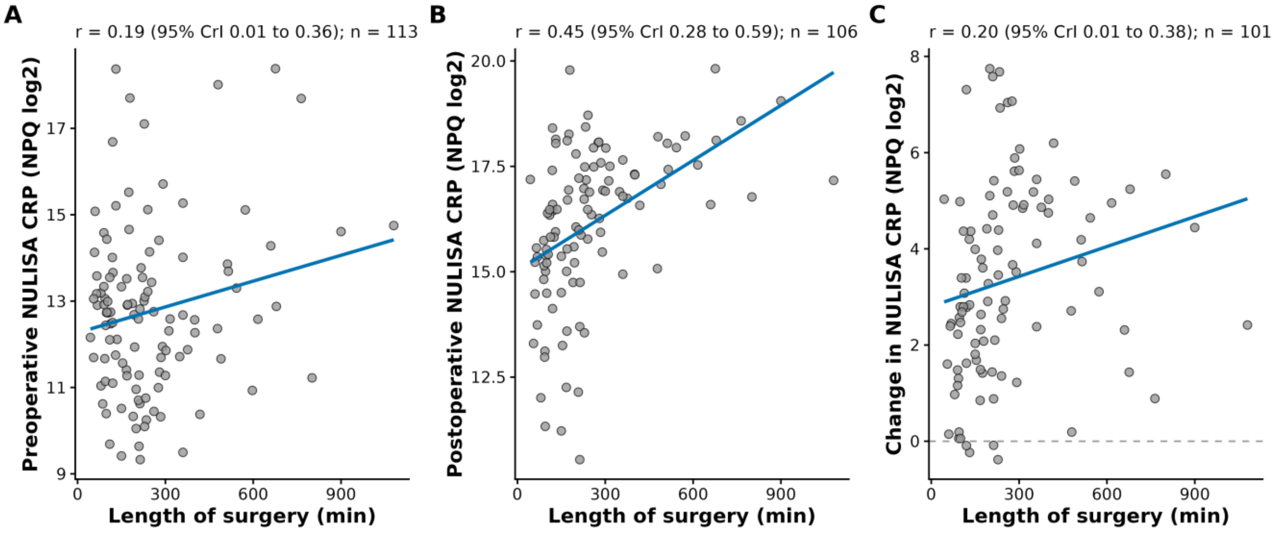


**(A)** Preoperative CRP, **(B)** postoperative day 1 CRP, and **(C)** change in CRP from preoperative to postoperative. NULISA CRP is shown in NQP units (log2 scale), so a change of 1 corresponding to a doubling of CRP; the dashed line in panel C indicates no change. Each point represents one patient; lines show linear fits. r is the posterior median correlation from a Bayesian bivariate normal model, with 95% credible interval (CrI). CRP = C-reactive protein; CrI = credible interval; NULISA = Nucleic acid Linked Immuno-Sandwich Assay; NPQ = NULISA protein quantification.

**Supplementary Table 1.** Characteristics of non-cardiac surgery patients by steroid group.

| **Characteristic** | **Dexamethasone (n = 56)** | **No steroid (n = 43)** | **Missing n (%)** |
|---|---|---|---|
| **Demographics** | | | |
| Age (years) | 69.0 (65.5, 73.5) | 70.0 (67.0, 76.0) | 0 (0%) |
| Male sex | 32 (57.1%) | 27 (62.8%) | 0 (0%) |
| BMI (kg $m^{-2}$) | 28.9 (25.4, 33.1) | 26.4 (23.4, 29.0) | 0 (0%) |
| **Medical history** | | | |
| Pre-existing atrial fibrillation | 4 (7.1%) | 4 (9.3%) | 0 (0%) |
| Existing memory concern | 7 (12.7%) | 8 (23.9%) | 9 (9.1%) |
| NSQIP 30-day mortality risk (%) | 0.5 (0.2, 1.2) | 0.8 (0.3, 1.8) | 0 (0%) |
| Baseline cognitive function | 32.0 (22.5, 37.0) | 29.0 (25.0, 34.0) | 6 (6.1%) |
| **Surgery** | | | |
| Length of surgery (min) | 197.5 (106.5, 388.0) | 170.0 (110.0, 253.0) | 0 (0%) |
| **Delirium** | | | |
| Postoperative delirium | 4 (7.1%) | 7 (16.3%) | 0 (0%) |
| Peak delirium severity (0–21) | 2.0 (1.0, 2.0) | 2.0 (0.0, 3.0) | 1 (1.0%) |
| **Biomarkers** | | | |
| CRP, preoperative (NPQ) | 12.5 (11.3, 13.6) | 12.9 (12.2, 14.1) | 7 (7.1%) |
| CRP, postoperative (NPQ) | 15.8 (14.1, 17.1) | 16.5 (15.4, 17.4) | 15 (15%) |

Values are median (IQR) or n (%). Non-cardiac surgery patients with known steroid status. Delirium was assessed using the 3D-CAM, except in two patients assessed using the CAM-ICU. Baseline cognitive function = sum of words recalled in two verbal fluency (COWAT) tests. BMI = body mass index; NSQIP = National Surgical Quality Improvement Program; CRP = C-Reactive Protein; NPQ = NULISA protein quantification (log2 scale).

**Supplementary Table 2.** Dexamethasone dose and postoperative NULISA CRP.

| Term | Estimate (95% CrI) | % change (95% CrI) | P(>0) |
|---|---|---|---|
| **Dexamethasone dose** | | | |
| 4 mg vs none | −0.61 (−1.60, 0.38) | −35% (−67%, 30%) | 0.113 |
| 8 mg vs none | −0.83 (−1.53, −0.14) | −44% (−65%, −10%) | 0.010 |
| 8 mg vs 4 mg | −0.22 (−1.26, 0.85) | −14% (−58%, 80%) | 0.338 |
| **Covariates** | | | |
| Preoperative CRP (per doubling) | 0.41 (0.26, 0.57) | 33% (19%, 48%) | >0.999 |
| Age (per 10 years) | −0.39 (−0.89, 0.12) | −24% (−46%, 8%) | 0.065 |
| Male sex | 0.30 (−0.36, 0.94) | 23% (−22%, 92%) | 0.815 |
| NSQIP mortality risk (per SD) | 0.13 (−0.22, 0.48) | 9% (−14%, 39%) | 0.766 |
| Length of surgery (per hour) | 0.21 (0.12, 0.30) | 16% (8%, 23%) | >0.999 |

Estimates are posterior median differences in postoperative day 1 NULISA CRP from Bayesian linear regression, shown on the NPQ (log2) scale and as percentage change in CRP, calculated as (2^estimate − 1) × 100; P(>0) is the posterior probability of a positive association (higher CRP), so the probability that a dose is associated with lower CRP is 1 − P(>0). Dexamethasone dose was entered as a categorical variable (none, 4 mg, 8 mg) with no dexamethasone as the reference; the 8 mg vs 4 mg comparison was derived from the same model. The model was adjusted for preoperative CRP (per NPQ unit, approximately a doubling of CRP), age (per 10 years), sex, NSQIP 30-day mortality risk (per SD) and length of surgery (per hour). n = 79 (none, n = 35; 4 mg, n = 10; 8 mg, n = 34); one patient who received 12 mg was excluded. The model was fitted with brms default priors. CrI, credible interval; CRP, C-reactive protein; NPQ, NULISA protein quantification; NSQIP, National Surgical Quality Improvement Program; SD, standard deviation.

**Supplementary Table 3.** Association between dexamethasone and postoperative delirium.

| Term | Unadjusted (n = 93)<br>OR (95% CrI)<br>P(OR>1) | Adjusted (n = 93)<br>OR (95% CrI)<br>P(OR>1) | Adj. + length of surgery (n = 93)<br>OR (95% CrI)<br>P(OR>1) |
|---|---|---|---|
| **Exposure** | | | |
| Dexamethasone (vs none) | 0.29 (0.06, 1.11)<br>0.038 | 0.29 (0.06, 1.20)<br>0.043 | 0.14 (0.02, 0.75)<br>0.013 |
| **Covariates** | | | |
| Age (per 10 years) | — | 1.00 (0.28, 3.27)<br>0.502 | 1.45 (0.32, 6.41)<br>0.687 |
| Male sex | — | 0.96 (0.22, 4.42)<br>0.479 | 0.97 (0.18, 5.53)<br>0.488 |
| NSQIP mortality risk (per SD) | — | 0.97 (0.40, 1.90)<br>0.471 | 0.79 (0.23, 1.76)<br>0.300 |
| Baseline cognition (per SD) | — | 0.76 (0.36, 1.60)<br>0.230 | 0.68 (0.26, 1.60)<br>0.192 |
| Length of surgery (per hour) | — | — | 1.45 (1.18, 1.87)<br>1.000 |

Estimates are posterior median odds ratios (OR) from Bayesian logistic regression, with 95% credible intervals (CrI); P(OR > 1) is the posterior probability that the odds of delirium are increased, so the probability that dexamethasone reduces the odds of delirium is 1 − P(OR > 1). The adjusted model includes age (per 10 years), sex, NSQIP 30-day mortality risk (per SD) and baseline cognitive function (per SD); the third model additionally includes length of surgery (per hour). Continuous covariates were standardised within this sample. All models included the same patients (n = 93, 10 with delirium; non-cardiac surgery patients with known steroid status and complete data). Given the small number of events, estimates, particularly from the third model, should be interpreted with caution. Models were fitted with brms default priors. — indicates the term was not included in the model. CrI, credible interval; NSQIP, National Surgical Quality Improvement Program; OR, odds ratio; SD, standard deviation.
